# CT perfusion stroke lesion threshold calibration between deconvolution algorithms

**DOI:** 10.1101/2022.11.17.22282418

**Authors:** Kevin J. Chung, Danny De Sarno, Ting-Yim Lee

**Affiliations:** Department of Medical Biophysics, University of Western Ontario, London, ON; Robarts Research Institute and Lawson Health Research Institute, London, ON; Department of Medical Imaging, University of Western Ontario, London, ON

**Keywords:** Acute ischemic stroke, cerebral blood flow, CT perfusion, stroke lesions, perfusion thresholds

## Abstract

**Objective:** CTP is an important diagnostic tool in managing patients with acute ischemic stroke, but challenges persist in the reliability of stroke lesion volumes determined with different software. We investigated a systematic method to calibrate CTP lesion thresholds between deconvolution algorithms using a digital perfusion phantom.

**Approach:** The accuracy of one model-independent and two model-based deconvolution algorithms in estimating ground truth cerebral blood flow (CBF) and Tmax in the phantom was quantified. Reference thresholds for ischemic core and penumbra were model-independent CBF<30% and Tmax>6 s, respectively, which is the current clinical standard. The equivalent model-based CBF and Tmax thresholds were determined by comparing linear regressions of phantom ground truth and deconvolution-estimated perfusion between algorithms. Calibrated thresholds were then validated in 63 patients with large vessel stroke by comparing admission CTP ischemic core and <3-hour diffusion-weighted imaging (DWI) lesion volume by Bland-Altman analysis. Agreement in target mismatch (core < 70 ml, penumbra ≥ 15 ml, mismatch ratio ≥ 1.8) determined by the three methods was assessed by Cohen’s kappa (κ) and concordance.

**Main Results:** The calibrated thresholds were CBF<15% and Tmax>6 s for both model-based methods. DWI minus CTP lesion mean volume differences (95% limits of agreement) were +16.2 (−30.9 to 63.3) ml, +10.9 (−32.9 to 54.7) ml, and +13.8 (−48.1 to 75.7) ml for model-independent and the two calibrated model-based approaches, respectively. Agreement in mismatch profiles with the two model-based deconvolution methods versus model-independent assessment was κ = 0.87 (95% confidence interval [CI]: 0.72 to 1.00) and κ = 0.86 (95% CI: 0.70 to 1.00), and both achieved 95% concordance.

**Significance:** We reported a systematic method of calibrating perfusion thresholds between deconvolution algorithms based on their quantitative accuracy. This may harmonize ischemic lesion volumes determined by different CTP software.

## Introduction

Imaging-based assessment of acute ischemic stroke, most often with computed tomography (CT), remains an important factor in evaluating eligibility for reperfusion treatment. Current guidelines recommend endovascular thrombectomy in patients with a demonstrated large vessel anterior circulation occlusion at CT angiography, and either mild-to-moderate early ischemic changes at non-contrast CT within 6 hours from stroke symptom onset or a favourable mismatch profile identified at CT perfusion (CTP) between 6 to 24 hours after stroke onset.^1,2^ A favourable mismatch is defined as a small ischemic core volume relative to the expected penumbra, which is assessed volumetrically by CTP,^3,4^ or by the National Institutes of Health Stroke Scale (NIHSS).^5^ Similarly, favourable CTP mismatch profiles were associated with better outcomes in patients treated with thrombolysis compared to placebo between 4.5 to 9 hours after stroke onset,^6^ which is beyond the <4.5-hour time window currently recommended in patients demonstrating mild-to-moderate early ischemic changes at non-contrast CT.^1^

Ischemic core and penumbral volumes are therefore diagnostic markers of interest in acute ischemic stroke. These markers are most often delineated with CTP parametric maps by identifying voxels above or below a pre-determined parameter threshold. In multiple clinical trials evaluating treatment eligibility with CTP,^3–6^ a single commercial CTP software (RAPID CTP, RapidAI, Menlo Park, CA) was used, which estimated ischemic core and penumbra by CBF<30% relative to the mean CBF in normally perfused brain and Tmax>6 s, respectively. These thresholds were validated in prior studies comparing CTP CBF lesions against near-concurrent diffusion-weighted MR imaging (DWI)^7^ and Tmax lesions against final infarct volume and clinical outcome.^8–10^ However, thresholds are specific to each CTP software, and variability in ischemic lesions determined by different CTP software packages have been reported.^11,12^ To demonstrate near-equivalence to RAPID CTP, software-specific optimal thresholds for ischemic core and penumbra have been re-derived, mainly by comparing against follow-up infarct volume, and were found to differ by software.^13–16^ These empirical threshold calibration techniques assume that follow-up infarct volume is roughly equivalent to true infarction at admission CTP, but infarct growth is expected in the time interval to follow-up imaging, even after successful endovascular thrombectomy. Empirical threshold calibration techniques are therefore dependent on the dataset and the extent to which follow-up infarct matches true admission infarct.

One key component of CTP software is the deconvolution algorithm used to estimate perfusion, which may differ in estimated perfusion between deconvolution methods. Kudo et al used a digital perfusion phantom in which CTP time-density curves (TDCs) were simulated to quantify CTP-estimated perfusion against the phantom’s ground truth perfusion.^17^ Differences in estimated perfusion were found between commercial CTP software, which incorporated different deconvolution algorithms. Since this study, the diagnostic endpoint of identifying ischemic core and penumbral volumes with CTP has been clearly defined and validated in clinical trials, and it is of renewed interest to characterize how choices in CTP algorithms affect the estimation of stroke lesion volumes. Specifically, it is desirable to determine a method to calibrate thresholds between deconvolution algorithms such that estimates of lesion volumes may be reproducible between CTP software. In this study, we showed that CTP ischemic core and penumbral thresholds could be calibrated between three deconvolution algorithms by comparing against ground truth perfusion from a digital perfusion phantom. This can harmonize ischemic lesion volumes determined by different CTP software.

## Method

### Theory

Concentration of contrast in tissue over time, *Q*(*t*), is related to that of a feeding artery by the governing equation:

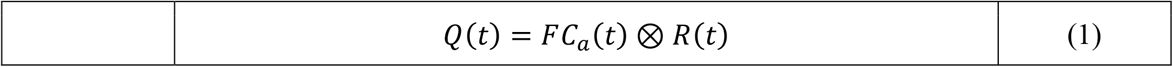

where *F* is the blood flow (in units of ml/min/100 g) delivering contrast to tissue, *C*_*a*_(*t*) is the concentration of contrast in the feeding artery as a function of time, ⨂ is the convolution operator, and *R*(*t*) is the impulse residue function (IRF), which describes the proportion of contrast remaining in tissue over time when a unit mass of contrast is injected as a short bolus into the tissue. A flow-scaled IRF, *R*_*F*_(*t*), is also commonly used and is defined as *R*_*F*_(*t*)= *FR(t*). The IRF is decoupled from the influence of the arterial input and describes the hemodynamic characteristics of the local tissue. As such, the perfusion parameters derived from the IRF are of diagnostic interest, and voxel-wise evaluation of the perfusion parameters result in quantitative maps. Perfusion parameters include the mean transit time (MTT) of contrast through the vasculature, which is the area underneath the IRF, and the Tmax, defined as the time-to-maximum of the IRF. Additionally, the cerebral blood volume (CBV) is related to CBF and MTT by the Central Volume Principle:^18^

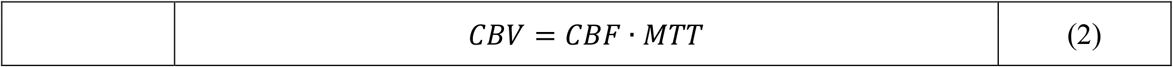

CBV is the volume of flowing blood in units of ml/100 g in the tissue vasculature. MTT and Tmax are normally expressed in units of seconds.

In a CTP study, the IRF cannot be directly measured from the native CT images, but rather, must be calculated by voxel-wise deconvolution of the arterial TDC, *C*_*a*_(*t*), from the measured tissue TDC, *Q*(*t*). Different methods for deconvolution have been proposed, which lead to differences in estimated perfusion and thus CTP stroke lesion thresholds.^17^ We compared CTP stroke lesion thresholds for three deconvolution algorithms: (1) Fourier Transform-based deconvolution,^19^ (2) model-based deconvolution with a plug flow model of contrast transport,^20,21^ and (3) model-based deconvolution with a Johnson-Wilson-Lee (JWL) model of contrast transport.^22^ The first two were implemented in-house as representative model-independent and model-dependent deconvolution methods. Of note, the model-independent deconvolution algorithm has been described in detail in a previous study^19^ and was the algorithm incorporated into clinically-validated RAPID CTP software, so its CTP lesion thresholds (ischemic core: CBF<30%; penumbra: Tmax>6 s) were selected as the reference on which the other two methods’ thresholds were calibrated. The third method was available in commercial CT Perfusion 4D (CTP4D Version 16.0-2.216, GE Healthcare, Waukesha, WI), which we henceforth refer as the “CTP4D” algorithm to avoid confusion with the plug-flow model-dependent deconvolution method. We briefly review the details of each model-independent and model-dependent deconvolution method, though the technical details of the CTP4D algorithm are unavailable.

### Model-independent Deconvolution

The Fourier Transform-based method leverages the convolution theorem, which states that a convolution in the time domain is equivalent to the product of frequency spectra:

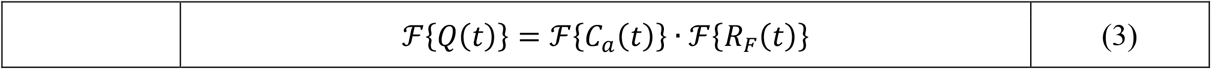

where ℱ{ ·} is the Fourier Transform. In numerical implementation, *Q*(*t*) and *C*_*a*_(*t*) are linearly interpolated to a virtual sampling interval of 1 s and zero-padded to twice their length to avoid time aliasing.^19^ This formulation is equivalent to circulant convolution in the time domain and therefore considered insensitive to delay in contrast arrival between the artery and the tissue (T0).^19,23^ The flow-scaled IRF can be estimated by the inverse Fourier Transform of the quotient between tissue and arterial TDC frequency spectrums. However, due to noise and other artifacts, the raw solution often results in flow-scaled IRFs with spurious oscillations that are not physiologically plausible. Fourier deconvolution is therefore regularized with a Wiener-like filter^19^ in the frequency domain using a 20% regularization threshold relative to the maximum magnitude of the arterial frequency spectrum. From the estimated flow-scaled IRF, CBF was determined as the peak height and Tmax as the time-to-maximum of the IRF. In the model-independent method, CBV was calculated as the area underneath the tissue TDC divided by the area underneath the arterial TDC. Areas were computed numerically by the trapezoidal rule. MTT was then calculated by the quotient of CBV and CBF as in the Central Volume Principle.^18^

### Model-dependent Deconvolution

The IRF of the model-dependent (plug-flow) method is given by:

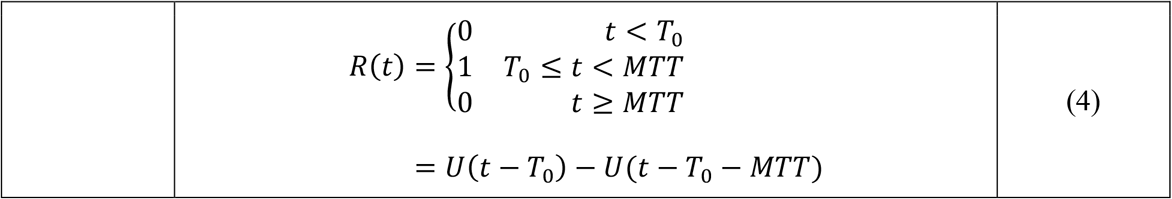

where T0 is the delay time between contrast arriving at the artery to arrival at tissue, MTT is the mean (uniform) transit time of contrast through the vasculature, and *U(t*) is the Heaviside unit step function. Substituting (4) into (1), a closed-form solution to *Q*(*t*) can be obtained:

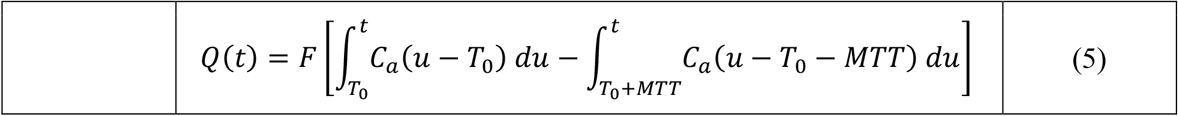

In other words, in the plug-flow model, the tissue TDC is the difference between the time-integral of the arterial TDC shifted by T0 and T0 + MTT, respectively, and linearly scaled by the CBF. We numerically solved (5) by a grid search of T0 and MTT, which linearizes the estimation of CBF. Specifically, all combinations of T0 ∈ [0, 1, 2 …, 23, 24] seconds and *MTT* ∈ [2, 3, …, 23, 24] seconds were searched, leading to a total of 575 grid search combinations per tissue TDC. A non-negative linear least squares algorithm^24^ solved for physiologically-constrained positive values of CBF for each T0 and MTT pair. The set of estimated parameters that produced the least squared difference between the measured and the estimated tissue TDC by (5) was considered the optimal parameters. In this method, T0 and MTT were directly estimated, and CBV was calculated by the Central Volume Principle in (2). Tmax was computed as Tmax = T0 + 0.5MTT, that is, time at half the width of the T0-shifted boxcar IRF.

While the implementation details of the CTP4D algorithm is unavailable, the IRF of the JWL model of contrast transport is:^22^

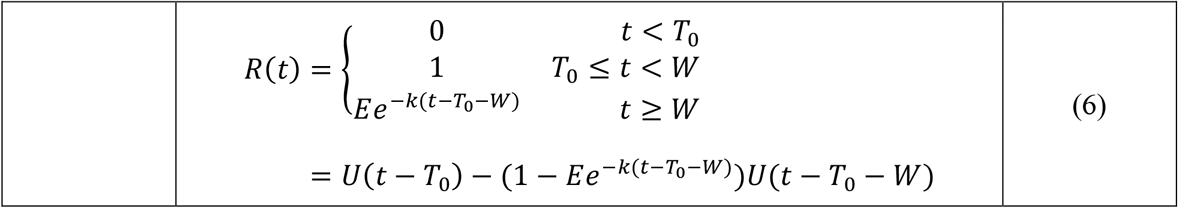

where *W* is the minimum transit time of contrast through the vasculature, *E* is the fraction of contrast with transit time > *W*, and *k* is a decay rate constant related to the heterogeneity of vascular transit time. Compared to the plug flow model, which assumes uniform transit time of contrast through vasculature, the JWL model appends an exponential decay following the boxcar IRF to account for heterogeneity in contrast transit time. MTT was the area underneath the IRF and Tmax was defined as Tmax = T0 + 0.5MTT.^16,22^

### Digital Perfusion Phantom-based Threshold Calibration

The quantitative accuracy of estimated perfusion parameters for each deconvolution algorithm was established by comparing against reference perfusion values in a digital perfusion phantom. Digital perfusion phantoms have been described previously^17^ and is briefly reviewed here. Tissue TDCs were simulated by convolving an assumed arterial TDC and simulated IRFs with known ground truth perfusion parameters (CBF, CBV, MTT, T0, and Tmax). Flow-scaled IRFs were simulated as gamma-variate functions to ensure that they had a different shape than the IRFs used in the model-based deconvolution methods. Gamma-variate IRFs were simulated with a wide range of ground truth perfusion parameters: *T*_0_ ∈ [0 0, 0.5, 1.0, 2.0, 3.0, 4.0, 8.0] s, *MTT* ∈ [3.4, 4.0, 6.0, 8.0, 10.0, 12.0, 16.0] s, and *CBV* ∈ [0.5, 1.0, 1.5, 2.0, 2.5, 3.0, 4.0, 5.0] ml/100 g. CBF was calculated as CBV/MTT by the Central Volume Principle;^18^ accordingly, CBF ranged from 1.9 to 88.2 ml/min/100 g at non-uniform intervals. Ground truth tissue TDCs were calculated by numerically convolving the simulated IRF and the linearly interpolated patient arterial curve at 0.01 s interval then resampled at 2 s interval. Zero-mean Gaussian noise with standard deviation, *σ* = 1.5 HU was added to the tissue TDCs to simulate the expected noise variation in tissue TDCs after standard Gaussian filtering of dynamic CTP images (Supplemental Figure 1). In total, 1024 noisy tissue TDCs were generated for each set of perfusion parameters by random sampling of Gaussian distributions with σ =1.5 HU. Further details on generating the phantom are in the Supplemental Materials.

We proposed to correlate deconvolution-estimated and ground truth phantom perfusion to calibrate stroke lesion thresholds between deconvolution methods. We focused on CBF and Tmax as they are most commonly used in practice to identify ischemic core and penumbra, respectively. First, the mean deconvolution-estimated CBF and Tmax over the 1024 noise realizations for each set of perfusion parameters was computed. Linear regressions between the deconvolution-estimated mean values and ground truth values were then calculated. These regression relationships were then used to construct calibration relationships for predicting the equivalent lesion thresholds between the three deconvolution thresholds. The derivation of the calibration relationship is provided in the Supplementary Materials. The equivalent model-based relative CBF threshold, *R*_*MB*_, to the reference model-independent relative CBF threshold, *R*_*MI*_, is given by:

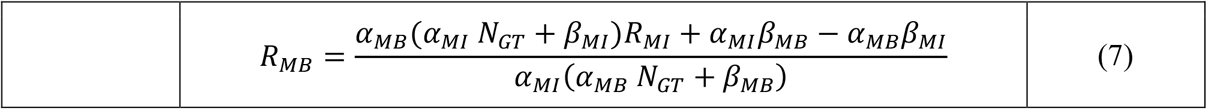

where *α* and *β* are linear regression slopes and intercepts, respectively, of model-independent (MI) and model-based (MB) deconvolution-estimated CBF against ground truth CBF in the digital perfusion phantom. *N*_*GT*_ = 50 ml/min/100 g and was the assumed ground truth CBF in the normally perfused brain for normalization of absolute CBF to relative CBF.^25^ Similarly, the calibration relationship between model-independent Tmax and model-based Tmax thresholds was:

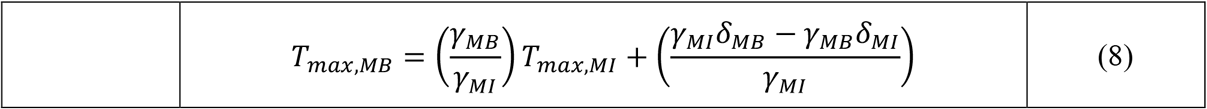

where *γ* and *δ* are linear regression slopes and intercepts, respectively, of model-independent and model-based deconvolution-estimated Tmax against ground truth Tmax in the digital perfusion phantom. The reference model-independent ischemic core and penumbra thresholds were *R*_*MI*_ = 30% and *T*_*max,MI*_ = 6 s, respectively.^4^

### Validation of Calibrated Threshold on Patient CTP Studies

The calibrated thresholds determined from the digital perfusion phantom experiments were validated in patient CTP studies. Validation data were from the ISLES 2018 Challenge,^26^ which consisted of 103 patients with a diagnosed acute large vessel occlusion within 8 hours of onset. CTP was performed at admission and diffusion-weighted MRI (DWI) within 3 hours of CTP. In a subset of 63 patients, DWI lesion segmentations were provided to serve as the reference for CTP ischemic core delineation. Only these 63 patients with reference DWI lesion volumes were included in our study; they were a subset of patients described in a previous publication,^7^ which was the main basis for the commonly used relative CBF<30% ischemic core threshold in RAPID CTP software.^5^ Patient demographics including age, NIHSS, onset-to-CT time, CT-to-DWI time, and CT scanner manufacturer for the entire 103-patient dataset were reported previously.^7,26^ Partitioned clinical data for the subgroup of 63 included patients of this study were not available. CTP studies were acquired with a wide range of scanners from different manufacturers, with different axial coverage (2-cm to 16-cm whole brain), and scan durations (43 to 64 s). All CTP studies were motion corrected and resampled to 1 s scan interval.^26^ Of note, patients with dual-slab CTP studies (required for short axial coverage scanners, denoted with a suffix “A/B” in the ISLES dataset) were processed as individual studies but grouped together as a single patient for statistical analysis.

CTP studies were automatically processed by an in-house Python-based 3D Slicer extension module for the model-independent and model-dependent deconvolution algorithms and the CTP4D software (Version 16.0-2.216, GE Healthcare, Waukesha, WI). The arterial and venous TDCs were automatically detected in a proximal healthy brain artery (e.g., middle or anterior cerebral artery or the basilar artery) and the sagittal sinus, respectively, and manually adjusted as required. In our in-house CTP software, skull-stripped dynamic CTP images were filtered by a 2D Gaussian kernel with a standard deviation of 2.4 mm. The model-independent and model-dependent deconvolution algorithms were then applied on identically pre-processed dynamic CTP studies to generate CBF, CBV, MTT, and Tmax maps. Similarly, CTP4D software pre-processed the dynamic images using a proprietary edge-preserving spatiotemporal filter and generated CBF, CBV, MTT, and Tmax maps using a JWL model-based deconvolution method.^22^

CTP4D software automatically segmented the ischemic core and penumbra using the determined calibrated CBF and Tmax thresholds. Uncalibrated threshold lesion volumes were also recorded for comparison. For our in-house software, the steps to segment the perfusion lesions were as follows. First, hypoperfused and normal tissue were considered parenchymal brain with Tmax greater than or lesser than or equal to 4 s, respectively. The mean CBF in the normally perfused brain was used as the reference CBF to calculate the relative CBF thresholds. Ischemic core was delineated within the hypoperfusion mask by model-independent CBF<30% and the calibrated model-dependent CBF threshold. Model-dependent CBF<30% lesions were also segmented for comparison. Similarly, the penumbra was segmented within the hypoperfused mask by model-independent Tmax>6 s and the calibrated model-dependent Tmax threshold. Hypoperfusion, ischemic core, and penumbra segmentations were post-processed by morphological operations (dilation, hole filling, then erosion using a 5-mm disk structure element) to improve robustness to CTP map noise. Of note, model-independent Tmax>6 s is an operational CTP definition of penumbra that may not reflect true penumbral tissue of intact but functionally inactive neurons.^27^ The mismatch ratio was calculated as the quotient of the penumbral volume and the ischemic core volume.

Correlation and agreement of ischemic core volume compared to DWI lesion volumes were assessed by the Pearson correlation coefficient and Bland-Altman analysis, respectively. Similarly, ischemic core and penumbral volumes between CTP software were compared by Pearson correlation and Bland-Altman analysis. Bland-Altman differences were reported as the reference (DWI or model-independent lesion volume) minus the compared technique. Patient-wise target mismatch profiles (core volume < 70 ml, penumbra volume ≥ 15 ml, mismatch ratio ≥ 1.8)^4^ were evaluated for each CTP software. Agreement between model-independent versus model-dependent, and model-independent versus CTP4D target mismatch profiles were assessed by Cohen’s kappa coefficient (κ). Cohen’s kappa was categorically scored as no agreement (κ < 0.00), slight (κ = 0.00 to 0.20), fair (κ = 0.21 to 0.40), moderate (κ = 0.41 to 0.60), substantial (κ = 0.61 to 0.80), and excellent (κ = 0.81 to 1.00) agreement.^28^ Accuracy, sensitivity, and specificity of target mismatch profiles were also computed using the model-independent determination as the reference. A two-tailed alpha < 0.05 was considered statistically significant.

## Results

### Threshold Calibration

Figure 1 summarizes the scatter plots and linear regressions of ground truth versus CTP-estimated CBF and Tmax. Model-independent and model-dependent deconvolution methods underestimated higher ground truth CBF, but underestimation was more severe with model-independent deconvolution. CTP4D CBF agreed well with ground truth CBF. Linear regression slopes were 0.36, 0.84, and 0.94 for model-independent, model-dependent, and CTP4D, respectively, and corresponding intercepts were 6.11, 4.12, and 5.98. The goodness of fits (*R*^2^) of linear regression were 0.76, 0.97, and 0.98 for model-independent, model-dependent, and CTP4D methods, respectively. Using the calibration relationship in Equation (7), the determined linear regression parameters, and *R*_*MI*_ = 30% as the reference threshold, the calibrated relative CBF thresholds were *R*_*MD*_ = 14.4% and *R*_4*D*_ *=* 16.6% for model-dependent and CTP4D methods, respectively, which were both rounded to 15% for validation in patient studies.

**Figure 1.**
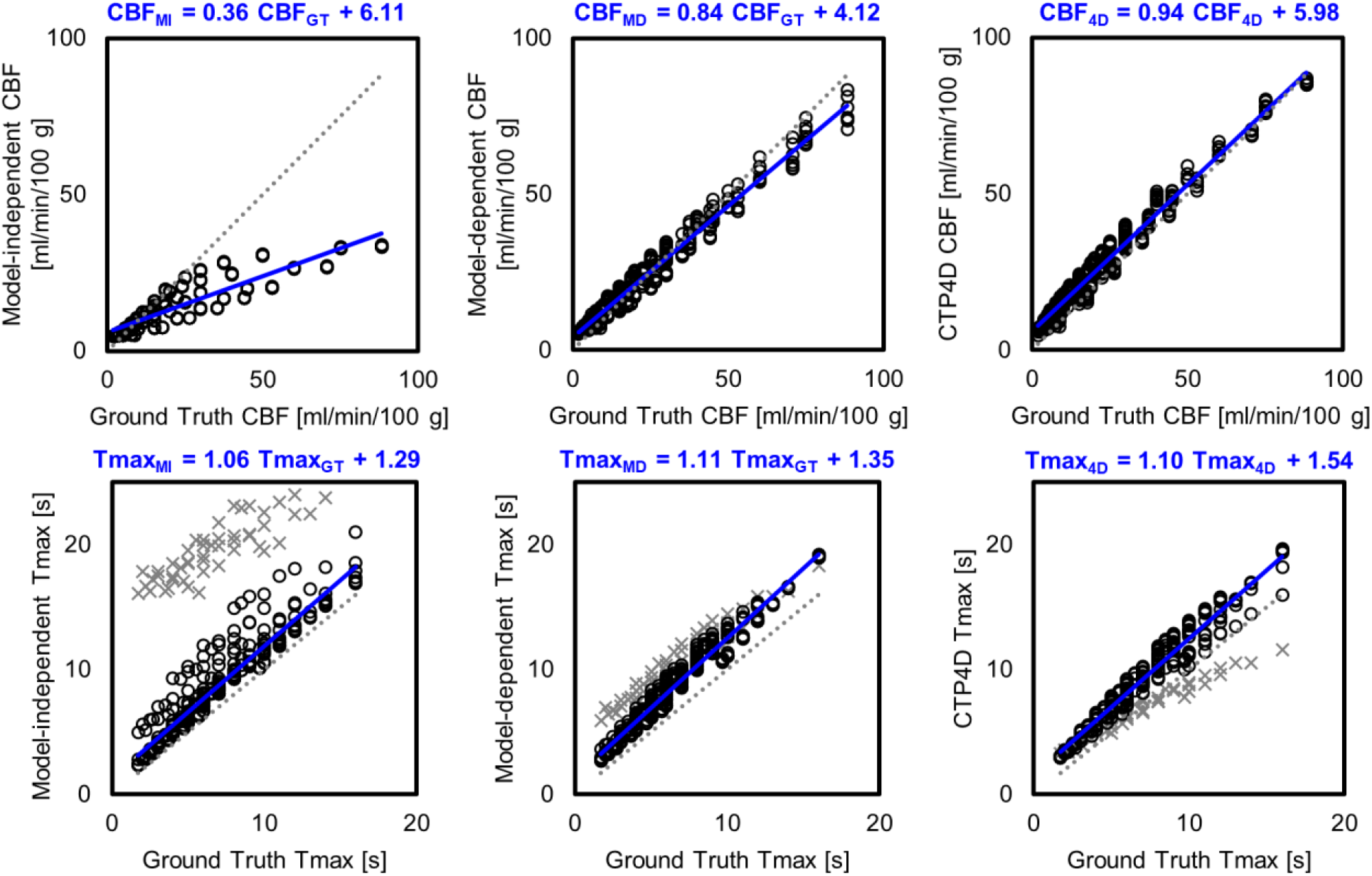
Ground truth versus cerebral blood flow (CBF; top row) and Tmax (bottom row) estimated by model-independent (MI; left), model-dependent (MD; middle), and CT Perfusion 4D (CTP4D) deconvolution. The dotted line indicates identity and linear regression is in blue. For Tmax, data points at cerebral blood volume of 0.5 ml/100 g were excluded from linear regression as those from the model-independent method deviated substantially from the majority due to low signal-to-noise ratio (grey crosses).

For Tmax threshold calibration, we found that model-independent Tmax substantially overestimated the ground truth at CBV of 0.5 ml/100 g due to low SNR (Figure 1, grey crosses). CTP-estimated Tmax values at CBV 0.5 ml/100 g were excluded for more robust linear regression against ground truth values. The linear regression slopes for ground truth versus model-independent, model-dependent, and CTP4D Tmax were 1.06, 1.11, and 1.10, and intercepts were 1.29 and 1.35, 1.54, respectively. Linear regression *R*^2^ were 0.89, 0.97, and 0.96 for model-independent, model-dependent, and CTP4D methods, respectively. Using the calibration relationship in Equation (8), the determined linear regression parameters, and *T*_*max,MI*_ *=* 6 s as the reference threshold, the calibrated Tmax thresholds were *T*_*max,MD*_ *=* 6.3 *s* and *T*_*max*,4*D*_ *=* 6.4 *s* for the model-dependent and CTP4D methods, respectively, which were rounded to 6 s for application on patient datasets.

### Validation of Calibrated Thresholds

Median (interquartile range, IQR) stroke lesion volumes from DWI and CTP are summarized in Table 1. Correlation plots and Bland-Altman analyses comparing DWI lesion versus CTP ischemic core volumes are shown in Figure 2. Plots for the uncalibrated CBF<30% ischemic core threshold are not shown as it clearly overestimated both DWI and model-independent relative CBF<30% lesion volumes (Table 1). Mean volume differences (95% limits of agreement) between DWI lesion versus model-independent CBF<30% and calibrated model-dependent CBF<15% and CTP4D CBF<15% were +16.2 (–30.9 to 63.3) ml, +10.9 (–32.9 to 54.7) ml, and +13.8 (–48.1 to 75.7) ml, respectively. Correlation and agreement of ischemic core and penumbra volumes estimated between CTP software was also strong (Figure 3) after threshold calibration, albeit model-independent lesion volumes were marginally smaller than model-dependent and CTP4D ones.

**Table 1.** Median (IQR) stroke lesion volumes from DWI, CBF, and Tmax with model-independent, model-dependent, and CTP4D software

| **Parameter** | **Median (IQR) Lesion Volume [ml]** |
| --- | --- |
| DWI Lesion | 47.6 (13.8 to 60.9) |
| Ischemic Core |  |
| Model-independent CBF<30% | 18.1 (8.9 to 38.3) |
| Model-dependent CBF<15% (calibrated) | 22.0 (12.2 to 44.2) |
| CTP4D CBF<15% (calibrated) | 22.4 (14.1 to 42.0) |
| Model-dependent CBF<30% (uncalibrated) | 71.7 (52.9 to 121.7) |
| CTP4D CBF<30% (uncalibrated) | 65.7 (40.0 to 100.6) |
| Ischemic Penumbra |  |
| Model-independent Tmax>6 s | 91.5 (57.4 to 149.3) |
| Model-dependent Tmax>6 s | 98.6 (69.3 to 157.8) |
| CTP4D Tmax>6 s | 89.3 (65.1 to 156.3) |

IQR indicates interquartile range; DWI, diffusion-weighted imaging; CBF, cerebral blood flow;
CTP4D, CT Perfusion 4D.

**Figure 2.**
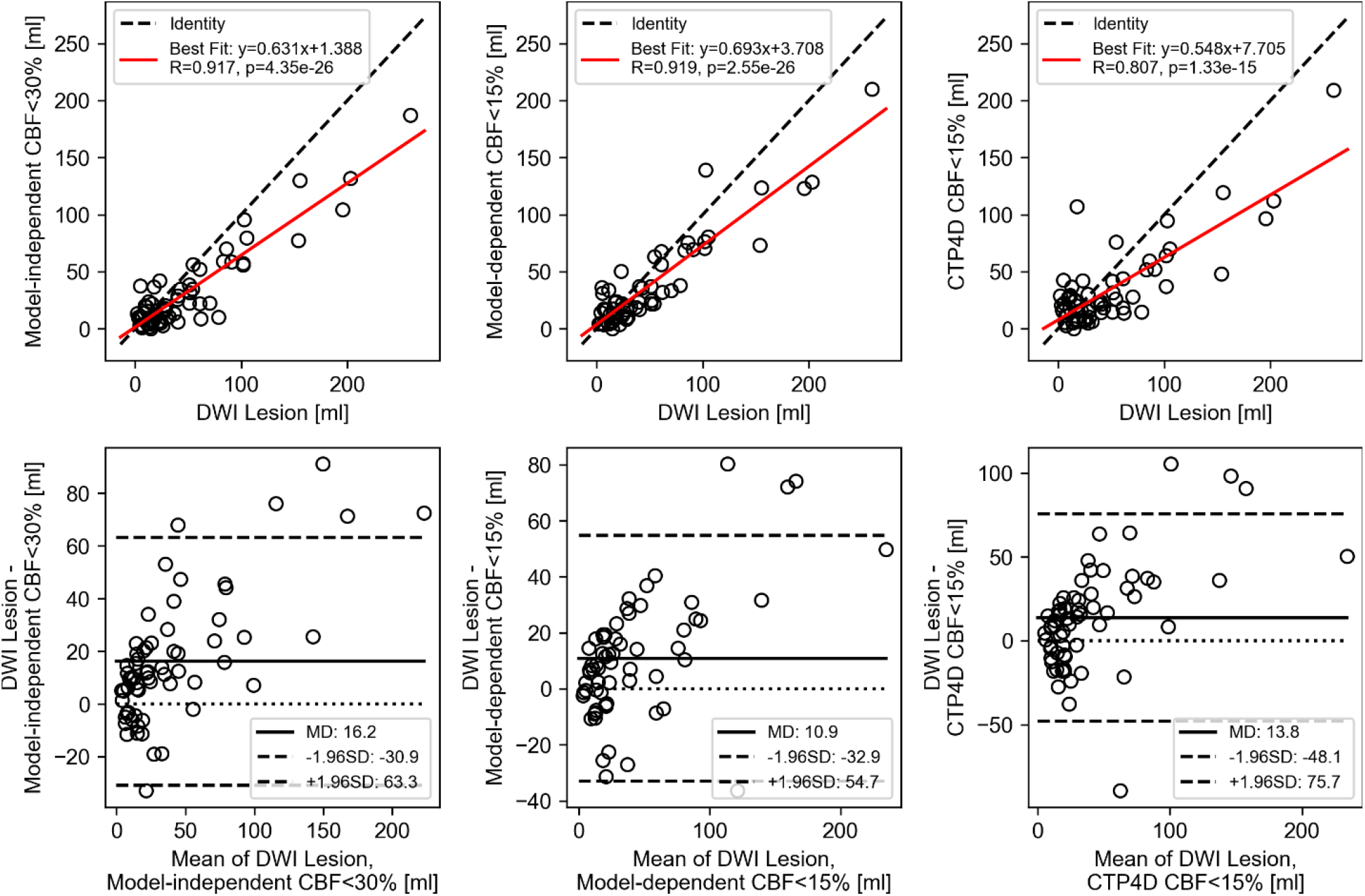
Correlation (top row) and Bland-Altman analysis (bottom row) of diffusion-weighted imaging (DWI) lesion volume versus model-independent cerebral blood flow (CBF)<30% (left column), model-dependent CBF<15% (middle column), and CT Perfusion 4D (CTP4D; right column) CBF<15%. The positive mean difference (MD) indicates underestimation of DWI lesion volume by all three CT perfusion software, likely due to infarct growth in the time interval between CT perfusion and DWI.

**Figure 3.**
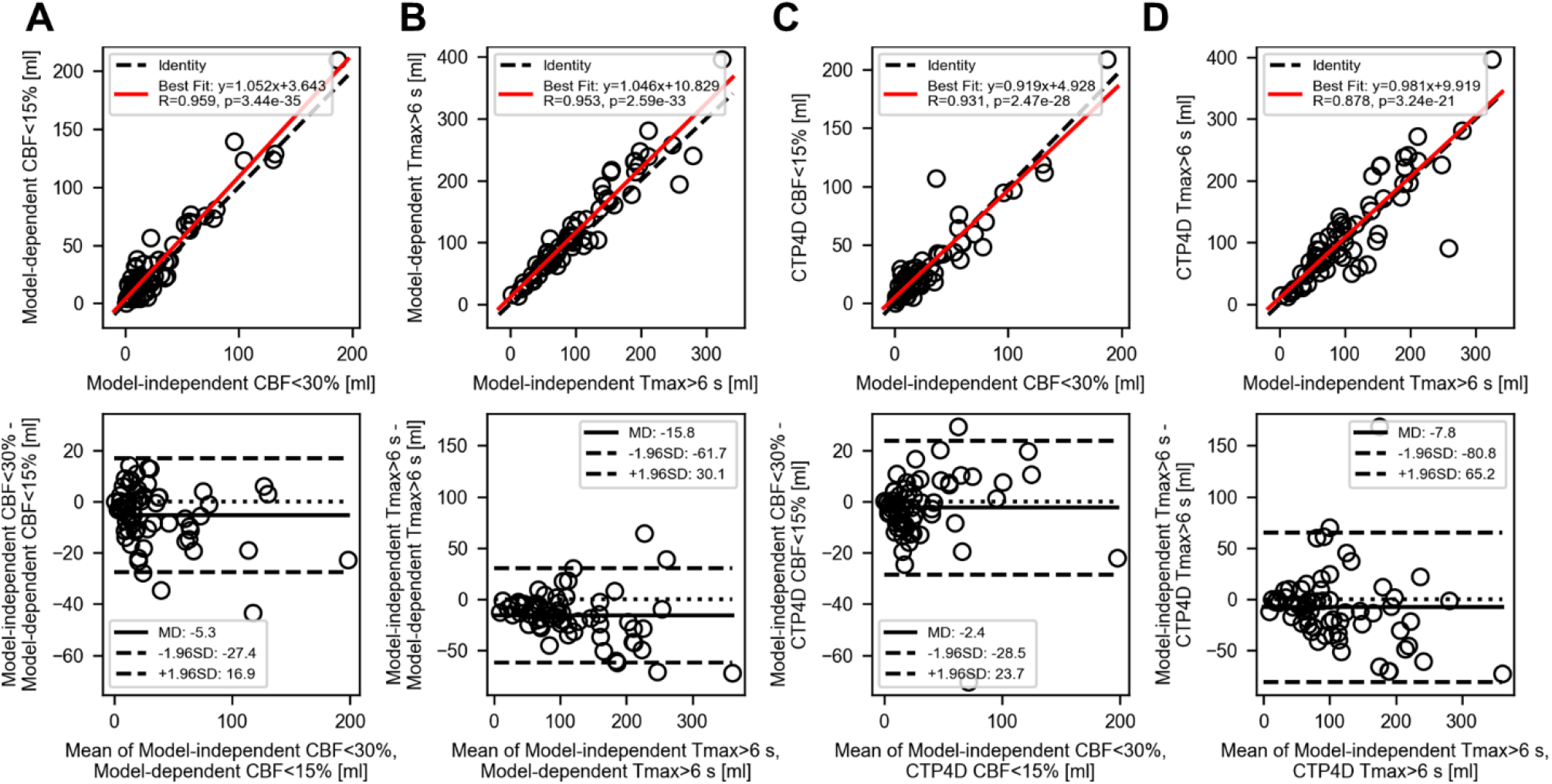
Correlation (top row) and Bland-Altman analysis (bottom row) of reference model-independent deconvolution lesion volumes versus (A, B) model-dependent and (C, D) CTP4D lesion volumes after threshold calibration. The reference ischemic core threshold was model-independent cerebral blood flow (CBF)<30% (A, C) and Tmax>6 s (B, D). The small negative mean differences (MD) indicate that model-independent lesion volumes were marginally overestimated, but nonetheless maintained strong correlation (Pearson correlation, R>0.87) between software.

Of the 63 included patients, 50 met the mismatch criteria and 13 did not by model-independent deconvolution assessment. Patients who did not meet the mismatch criteria by model-independent deconvolution had a core volume > 70 ml (N=4), mismatch ratio < 1.8 (N=3), or a combination of negative criteria (N=6). Agreement and classification metrics between CTP software using model-independent target mismatch profile as the reference are summarized in Table 2, including subgroup analyses for core volume, penumbral volume, and mismatch ratio. Overall, excellent agreement was found in target mismatch profiles determined from model-independent deconvolution and model-dependent deconvolution (κ = 0.87, 95% confidence interval [CI]: 0.72 to 1.00) and CTP4D software (κ = 0.86, 95% CI: 0.70 to 1.00). Strong agreement was also found in classifying favourable/unfavourable core and penumbra volumes but weaker for mismatch ratios (Table 2). Sensitivity and accuracy of classifying a favourable/unfavourable stroke lesion profile was overall excellent. Figure 4 and Figure 5 show examples of concordant and discordant target mismatch profiles between CTP software.

**Table 2.** Sensitivity, specificity, accuracy, and Cohen’s kappa (κ) of classifying target mismatch criteria with model-independent deconvolution versus model-dependent deconvolution and CT Perfusion 4D (CTP4D) software

| Criteria | Sensitivity (%) | Specificity (%) | Accuracy (%) | $\kappa$ (95% CI) |
| --- | --- | --- | --- | --- |
| Model-dependent |  |  |  |  |
| Target Mismatch Profile | 47/50 (94) | 13/13 (71) | 60/63 (95) | 0.87<br>(0.72 to 1.00) |
| Core < 70 ml | 53/55 (96) | 8/8 (100) | 61/63 (97) | 0.87<br>(0.70 to 1.00) |
| Penumbra ≥ 15 ml | 61/61 (100) | 2/2 (100) | 63/63 (100) | 1.00<br>(1.00 to 1.00) |
| Mismatch Ratio ≥ 1.8 | 52/54 (96) | 5/9 (56) | 57/63 (90) | 0.57<br>(0.27 to 0.88) |
| CT Perfusion 4D |  |  |  |  |
| Target Mismatch Profile | 48/50 (96) | 12/13 (92) | 60/63 (95) | 0.86<br>(0.70 to 1.00) |
| Core < 70 ml | 53/55 (96) | 6/8 (75) | 59/63 (94) | 0.71<br>(0.45 to 0.98) |
| Penumbra ≥ 15 ml | 61/61 (100) | 2/2 (100) | 63/63 (100) | 1.00<br>(1.00 to 1.00) |
| Mismatch Ratio ≥ 1.8 | 53/54 (98) | 5/9 (56) | 58/63 (92) | 0.62<br>(0.33 to 0.92) |

**Figure 4.**
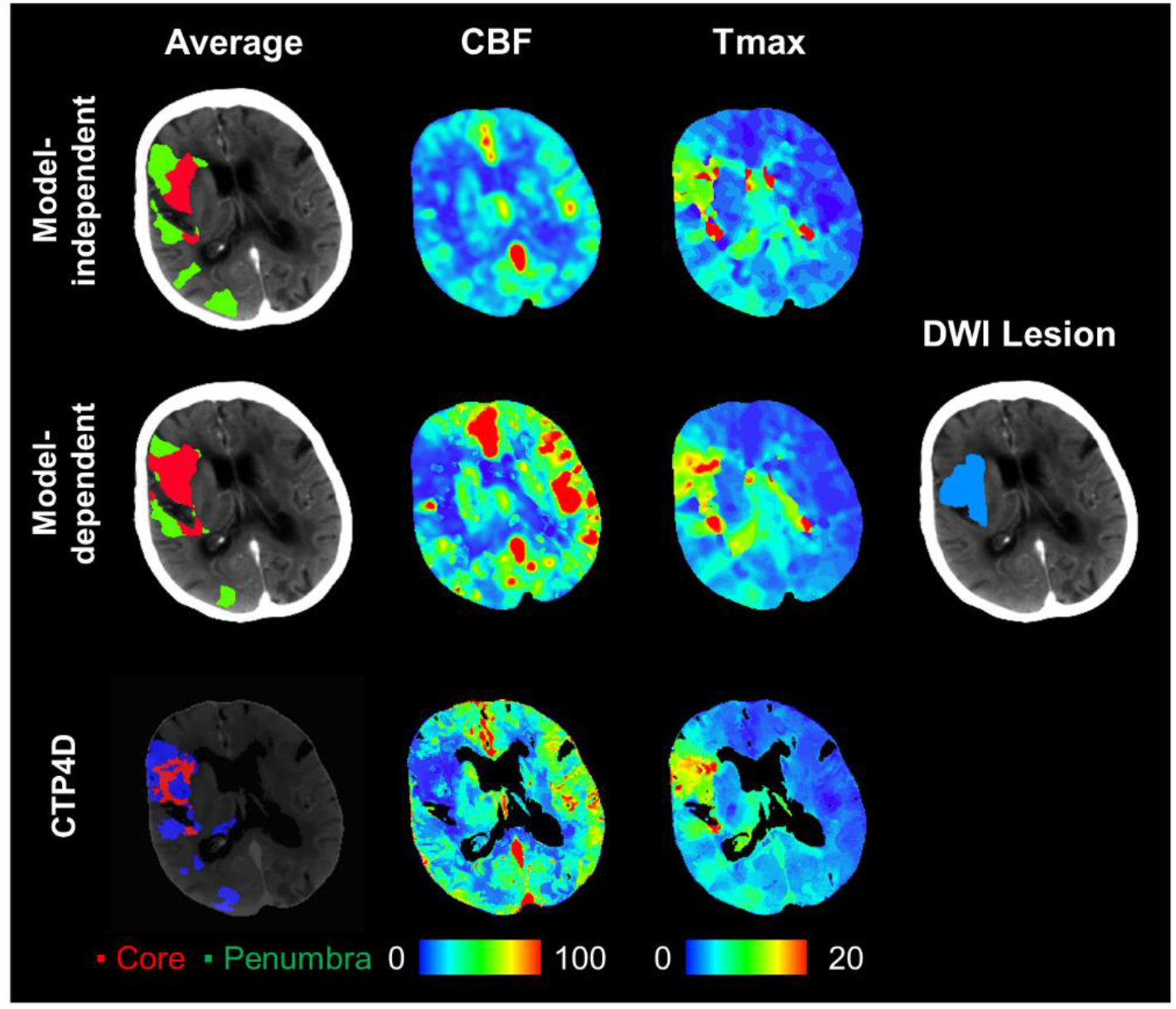
Summary of ischemic core (red) and penumbral segmentations (green/blue), cerebral blood flow (CBF) maps, Tmax maps, and DWI lesion segmentation (light blue). Model-independent and CTP4D software agreed in predicting a favourable mismatch profile, but the model-dependent approach predicted an unfavourable mismatch profile. Core volume (red), penumbral volume (green/blue), and mismatch ratio respectively were 26.6 ml, 68.8 ml, and 2.6 with the model-independent approach, and 37.3 ml, 62.3 ml, and 1.7 with the calibrated model-dependent thresholds, and 21.6 ml, 56.5 ml, and 2.6 with calibrated CTP4D thresholds. Diffusion-weighted imaging at <3-hour interval time showed a lesion volume of 40.2 ml (light blue). CBF is in units of ml/min/100 g and Tmax in seconds.

**Figure 5.**
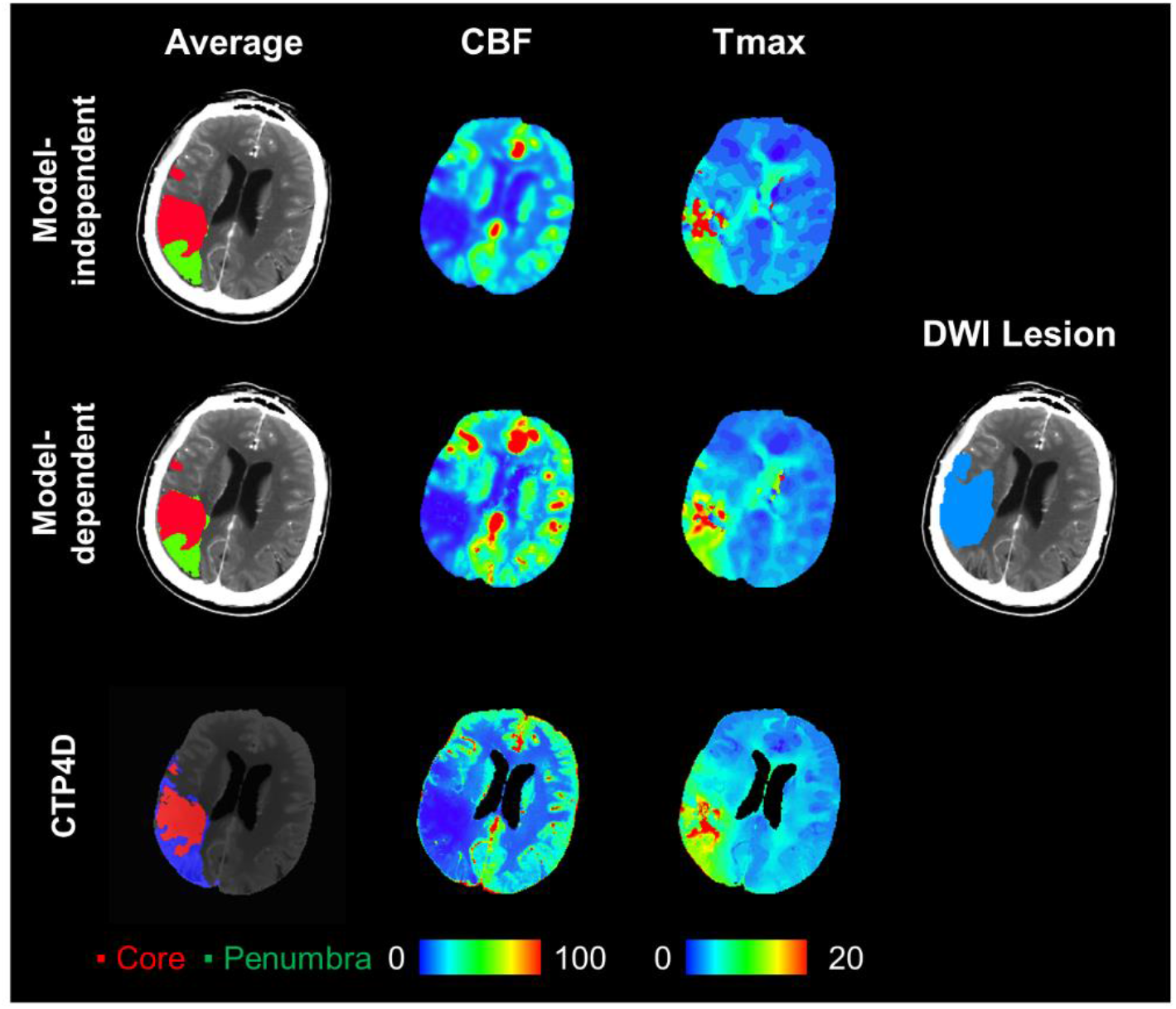
Summary of ischemic core (red) and penumbral segmentations (green/blue), cerebral blood flow (CBF) maps, Tmax maps, and DWI lesion segmentation (light blue). Model-independent and model-dependent approaches concorded in predicting an unfavourable mismatch profile, but CTP4D predicted a favourable mismatch profile, which may potentially encourage treatment based on imaging-based guidelines. Core volume (red), penumbral volume (green/blue), and mismatch ratio respectively were 25.8 ml, 35.2 ml, and 1.4 with the model-independent approach, and 23.5 ml, 37.8 ml, and 1.6 with the calibrated model-dependent thresholds, and 23.9 ml, 50.4 ml, and 2.1 with calibrated thresholds on CTP4D software. Diffusion-weighted imaging at <3-hour interval time showed a lesion volume of 39.6 ml (light blue). CBF is in units of ml/min/100 g and Tmax in seconds.

## Discussion

CTP is an important diagnostic tool in managing patients with acute ischemic stroke, but challenges persist in the reliability of stroke lesion volumes determined with different software packages.^11^ In this study, we reported a digital perfusion phantom-based method to systematically calibrate CTP lesion thresholds with respect to the deconvolution algorithm used. We found that model-independent deconvolution substantially underestimated higher ground truth CBF in the perfusion phantom, whereas the two model-based methods (plug flow^20^ and JWL^22^ models) had better agreement with ground truth CBF. The ischemic core CBF threshold for the two model-based methods therefore required reduction to CBF<15% with guidance from the perfusion phantom to match that of the model-independent CBF<30% threshold. Tmax penumbral thresholds were roughly similar between all deconvolution methods in our study. In patient CTP studies using DWI lesion volume as reference, we found ischemic core volume estimation was substantially improved after threshold calibration. Importantly, our phantom-based threshold calibration method was valid with CTP4D software, which used proprietary processing steps different to those used in our in-house CTP software, but nonetheless achieved strong agreement to reference (DWI) stroke lesion volumes and predicted mismatch profiles. Our novel contribution was to leverage quantitative relationships between deconvolution-estimated and ground truth perfusion in digital perfusion phantoms to calibrate perfusion thresholds between deconvolution methods.

Linear regression to quantify the parameter estimation accuracy has been reported previously,^17^ but has not been extended to systematically calibrate lesion thresholds between deconvolution methods. Previous threshold calibration studies empirically adjusted CTP lesion thresholds and demonstrated equivalence in estimating lesion volume to other validated CTP software.^13–15^ Whereas this empirical approach is likely dataset dependent, our phantom-based calibration technique is systematic and based on the quantification accuracy of each deconvolution algorithm. As confirmed by our digital perfusion phantom experiment, different deconvolution algorithms can have substantially different accuracy in estimating CBF, and consequently, CBF thresholds for ischemic core must be adjusted. This finding was supported by additional validation on patient CTP studies. In our study, the model-dependent method and CTP4D software, the latter which also uses model-based deconvolution, achieved roughly the same calibrated thresholds. This result does not necessarily imply that all model-based deconvolution methods have the same stroke lesion thresholds. The phantom-based calibration experiment should be performed for each CTP software, whether it uses deconvolution or not. Importantly, the commonly used relative CBF<30% threshold for ischemic core^7,29,30^ does not appear to be an absolute pathophysiological marker of infarction, but rather, is dependent on the deconvolution method used to estimate CBF. This may explain the well-known problem of widely different estimates of ischemic core and mismatch profiles from CTP maps generated by different software when a single relative CBF <30% threshold was used.^15,31^ In our analysis, the Tmax threshold did not have to be adjusted among the three deconvolution methods. This may be in part due to the simulated gamma-variate IRF was “forced” to have Tmax = T0 + 0.5MTT, which was the definition of Tmax in IRFs used in the two model-based deconvolution methods. Patient IRFs may have a different relationship between Tmax, T0, and MTT. Nonetheless, all three CTP software produced similar penumbral volumes in the patient CTP studies.

Target mismatch profile is of the greatest diagnostic interest in acute ischemic stroke CTP. In our study, substantial agreement in identifying favourable/unfavourable mismatch profiles was achieved between the three CTP software after threshold calibration. As shown by subcategory classification metrics (core < 70 ml, penumbra ≥ 15 ml, and mismatch ratio ≥ 1.8), disagreement was mostly in detecting favourable/unfavourable mismatch ratios. Mismatch ratio is the quotient of penumbral and core volumes, so the error in each measurement propagates to the mismatch ratio and worsens agreement between software. However, these incremental differences may not be as severe in clinical practice because a clinician may be able to rule out artefacts, integrate critical information from clinical assessments, and holistically make judgment calls when mismatch profiles are at the borderline between favourable and unfavourable. The few cases with discordant mismatch profiles may be due to differences between CTP software that cannot be accounted by our proposed linear calibration method. These differences may include the dependence of each deconvolution algorithm on T0, MTT, and signal-to-noise ratio, or other algorithmic differences such as dynamic image filtering and lesion segmentation methods. Though our phantom-based technique was successful in calibrating the lesion threshold and improving diagnostic agreement in most cases, further work is required to reduce discrepancies in stroke lesion volumes and mismatch profiles caused by these other CTP processing steps.

The validity of the threshold calibration technique is contingent on how well the phantom reproduces conditions expected in patient CTP studies. Each component of the phantom can be scrutinized to evaluate its fidelity to patient CTP studies. First, the assumed arterial curve was taken from a patient CTP study, which contained a small amount of noise relative to the large arterial signal. Arterial TDC noise was therefore incorporated into the convolved tissue TDCs, which may cause the simulated tissue TDC to be noisier than the real ones. We expect the small amount of added noise to be of little consequence. Further, the patient arterial TDC used was an improvement on prior digital phantoms that used an analytical gamma-variate arterial curve^17^ because it included contrast recirculation. Second, only gamma-variate IRFs were modeled in our phantom. A prior study showed that estimation of perfusion parameters with both model-independent and model-dependent deconvolution algorithms are dependent on the IRF model.^17^ As such, the calibration curve may depend on the selected IRF model for generating the digital perfusion phantom. However, canonical IRF shapes may vary by tissue subtype and status, such as normal/ischemic grey and white matter. Incorporating these factors into a digital perfusion phantom would be challenging. The gamma-variate IRF model was therefore chosen for practicality and to be different from the IRFs used in the model-dependent deconvolution methods. Third, we simulated a wider range of ground truth perfusion parameters in the digital perfusion phantom compared to prior studies^17,21^ to better balance the prevalence of low and high ground truth CBF and CBV in linear regression. Lastly, the injected Gaussian noise level of *σ* = 1.5 HU appears low compared to other studies,^17,21^ but this noise level was chosen to match that of dynamic CTP images after standard Gaussian filtering at the strength used in our study (Supplemental Figure 1). A fixed noise level, as opposed to one adjusted to meet a prescribed SNR,^17^ is required to simulate realistic conditions in which low-CBV tissue would have lower SNR than that of normal tissue. By injecting a post-filtering noise level, the need to filter the dynamic phantom images was also obviated. Gaussian noise added to the digital perfusion phantom is spatially uncorrelated, so Gaussian filtering would be more effective in the phantom compared to that in patient CTP studies where noise is spatially correlated.

This study had limitations. First, we did not have access to RAPID CTP software and its predicted stroke lesion volumes and mismatch profiles. Instead, reference stroke lesion volumes and target mismatch profiles were generated with our in-house implementation of the RAPID CTP model-independent method.^19^ However, we validated our ischemic core volume estimation against acute DWI, which achieved similar agreement to a previous validation study by Cereda et al in which Bland-Altman mean difference (95% limits of agreement) between DWI versus RAPID CTP CBF<30% was +12.0 (–25.9 to 49.2) ml.^7^ Our study was not a definitive comparison against RAPID CTP software, but rather, a demonstration that better agreement in mismatch profiles can be achieved between CTP software by using our proposed phantom-based threshold calibration method. Direct comparison against RAPID CTP should be performed in future studies to determine calibrated thresholds equivalent to RAPID CTP ischemic core and penumbral thresholds. Second, as with previous studies validating CTP ischemic core thresholds, the DWI lesion volume used as the reference was not the true infarct volume at the time of CTP. Sources of error include infarct growth in the time interval between CTP and DWI, co-registration errors, manual segmentation errors, and potential DWI lesion reversal,^32^ which could not be confirmed due to the absence of additional delayed imaging in this dataset.^7,26^ Furthermore, there was no ground truth for penumbral lesions; as such, only the agreement of Tmax>6 s lesions between deconvolution algorithms could be assessed. It remains to be determined whether lesion volumes from the calibrated threshold can correctly identify stroke treatment candidates and are associated with clinical outcomes. Additional validation on a large cohort of patients with acute ischemic stroke is warranted. Lastly, the median time from stroke onset to CT in our dataset was 185 (IQR: 180 to 238) min, which was outside the 6 to 24-hour CTP time window indicated by common best practice guidelines.^1^ Other guidelines do not discourage CTP in the early time window,^2^ and there are studies that suggest its benefit over standard neuroimaging alone.^33,34^ Our calibration method nonetheless requires additional validation in patients presenting in the 6 to 24-hour time window.

## Conclusion

We investigated a method to systematically calibrate CTP thresholds for the estimation of ischemic core and penumbral thresholds based on CBF and Tmax parametric maps generated by different deconvolution software. The quantitative accuracy of estimating perfusion parameters for each deconvolution algorithm was benchmarked by a digital perfusion phantom, and these relationships were used to perform inter-software perfusion parameter threshold calibration. Validation on patient CTP studies suggested that the calibrated thresholds substantially standardize stroke lesion volume estimation compared to uncalibrated thresholds. As such, diagnostic agreement between CTP software was substantially improved after threshold calibration as assessed by target mismatch profiles. To further improve diagnostic consistency between CTP software, future studies are warranted to better characterize how choices of pre- and post-deconvolution processing algorithms affect the detection of diagnostic markers in acute ischemic stroke.

## Data Availability

All data produced in the present study are available upon reasonable request to the authors

## Acknowledgments

The authors acknowledge the organizers of the ISLES 2018 Challenge for access to the invaluable imaging dataset.

## Supplemental Materials

### Generating the Digital Perfusion Phantom

A digital perfusion phantom uses Equation (1) to simulate tissue TDCs from an assumed arterial TDC and simulated IRFs with known ground truth parameters. The quantitative accuracy of a deconvolution algorithm can then be benchmarked by deconvolving the arterial TDC from the simulated TDC to estimate perfusion parameters and compare against the known ground truth values.

In our digital perfusion phantom, the arterial TDC was taken from the internal carotid artery in the healthy brain hemisphere of an acute ischemic stroke CT perfusion (CTP) study with a uniform image interval of 1.8 s over 81 s. No further filtering or curve fitting was applied. Flow-scaled IRFs were simulated as gamma-variate functions to ensure that they had a different shape than the IRFs used in the model-based deconvolution methods. Gamma-variate IRFs were simulated with a wide range of ground truth perfusion parameters: *T*_0_ ∈ [0.0, 0.5, 1.0, 2.0, 3.0, 4.0, 8.0] s, *MTT* ∈ [3.4, 4.0, 6.0, 8.0, 10.0, 12.0, 16.0] s, and *CBV* ∈ [0.5, 1.0, 1.5, 2.0, 2.5, 3.0, 4.0, 5.0] ml/100 g. CBF was calculated as CBV/MTT by the Central Volume Principle;^18^ accordingly, CBF ranged from 1.9 to 88.2 ml/min/100 g at non-uniform intervals. Ground truth tissue TDCs were calculated by numerically convolving the simulated IRF and the linearly interpolated patient arterial curve at 0.01 s interval then resampled at 2 s interval. Zero-mean Gaussian noise with standard deviation, *σ =* 1.5 HU was added to the tissue TDCs to simulate the expected noise variation in tissue TDCs after standard Gaussian filtering of dynamic CTP images (Supplemental Figure 1). In total, 1024 noisy tissue TDCs were generated for each set of perfusion parameters by random sampling of Gaussian distributions with σ =1.5 HU. Partial volume effect, hematocrit ratio, and tissue density were neglected or set to 1.

Simulated noisy tissue TDCs were arranged in a square pattern as in previous studies^17,21^ and as illustrated in Supplemental Figure 2 by the ground truth perfusion values. Briefly, the 1024 noisy tissue curves with the same ground truth perfusion parameter were arranged in a 32 × 32 tile. Tiles were then arranged in a 7 × 7 grid according to their ground truth *T*_0_ and MTT. MTT varied by column (longest MTT in the leftmost column, shortest MTT on the rightmost column) and *T*_0_ varied by row (shortest on the topmost row, longest on the bottommost row). CBV was constant for each 7 × 7 grid but varied over the slice axis. CBF increased from left to right (long to short MTT) and varied according to the CBV in each slice. The described procedure generated a simulated CTP study of 8 slices and 40 time points over 80 s for each slice. The dynamic images of the digital perfusion phantom were saved as DICOM files using a rescale slope of 0.1 to maintain voxel value precision up to a tenth of a decimal.

Each CTP software (model-independent deconvolution, model-dependent deconvolution, and CTP4D) processed the digital perfusion phantom independently using the original arterial TDC and performed voxel-wise deconvolution of each tissue TDC. The software did not filter the dynamic CTP images because the injected noise was already at levels expected after filtering. To match the generation of the phantom, partial volume effect, hematocrit ratio, and tissue density were neglected or set to 1.

**Supplemental Figure 1.**
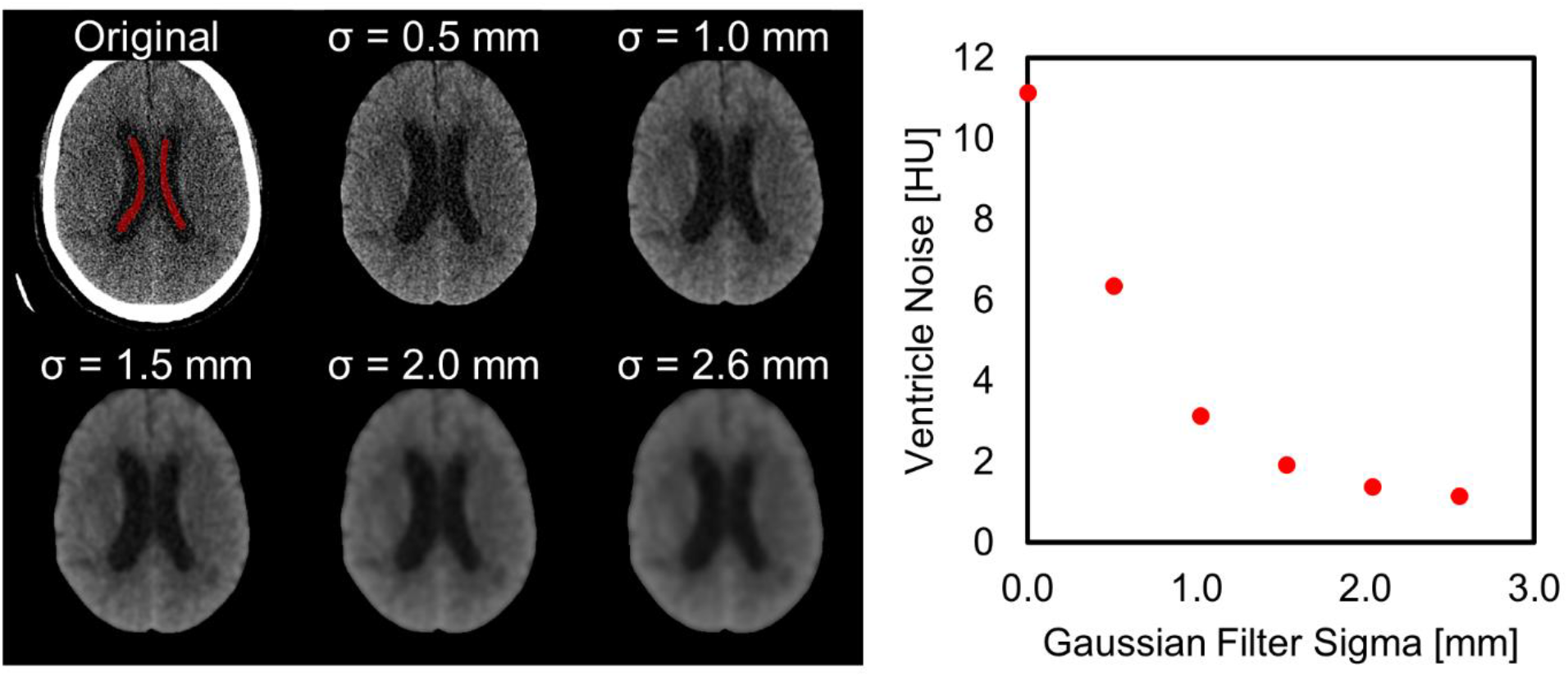
Noise level expected in dynamic CT perfusion images after filtering with Gaussian kernels of different strengths (*σ*). Noise was estimated in a uniform intracranial region, i.e., the brain ventricles, as indicated by the red segmentation in the original unfiltered image. The dynamic CT perfusion study shown was acquired using a standard brain CT perfusion protocol (tube voltage: 80 kV, tube current-exposure time product: 100 mAs, slice thickness: 5 mm, 44 dynamic images over 80 s at uniform intervals). Noise was the standard deviation of delineated ventricle voxels over all 44 dynamic images. The ventricle noise at the Gaussian filter strength used in this study (*σ =* 2.4 mm) was approximately 1.5 HU.

**Supplemental Figure 2.**
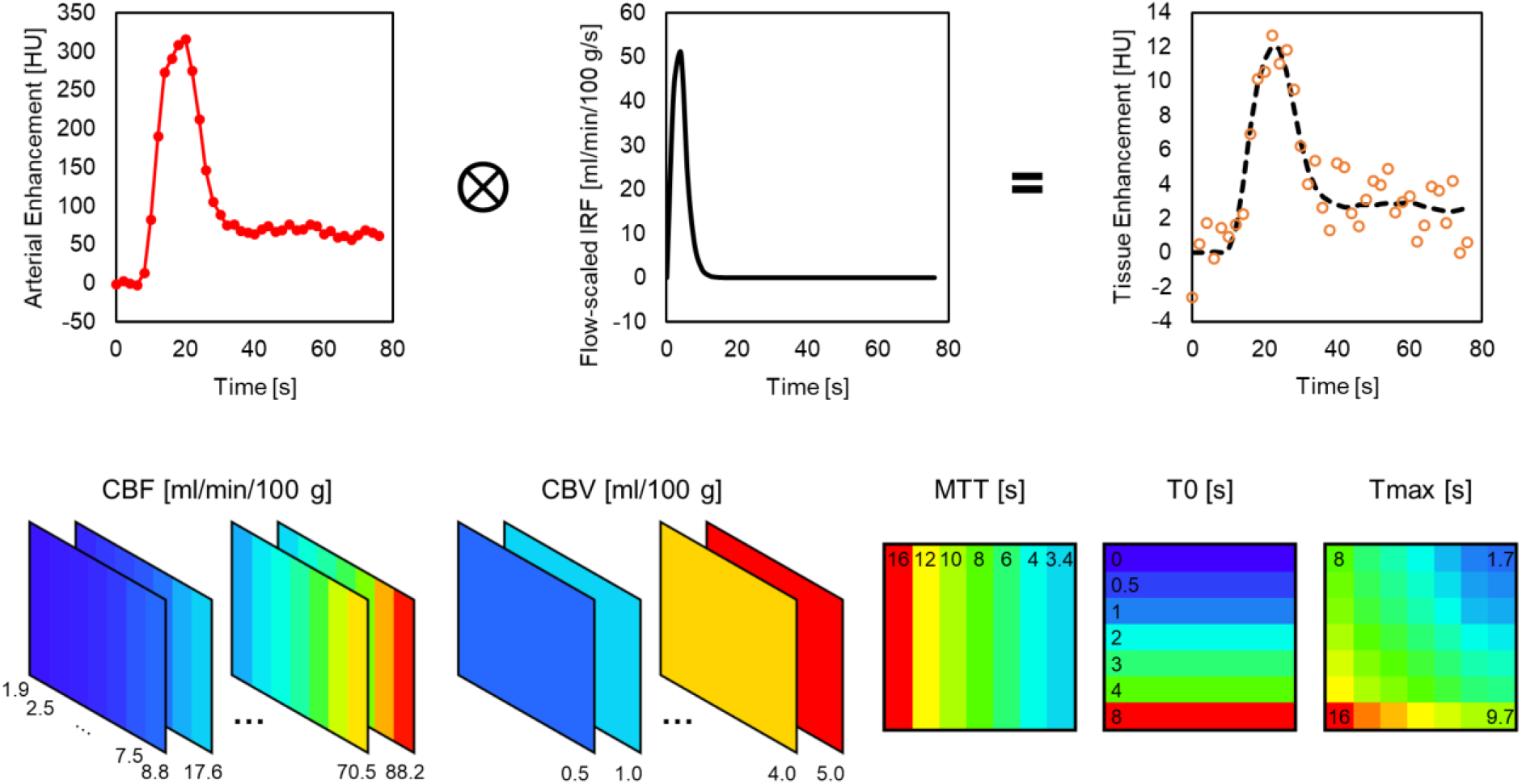
Generating the digital perfusion phantom. The arterial curve (top left) was convolved (⨂) with a gamma-variate flow-scaled impulse residue function (top centre) to produce a ground truth tissue curve (top right, dotted line). Gaussian noise (*σ =* 1.5 HU) was added to the simulated tissue curve. Each set of simulated perfusion parameters had a 32 × 32 tile of tissue time-density curves each with a different noise realization. Tiles were arranged in a 7 × 7 pattern as in the ground truth perfusion phantom shown in the bottom row. Cerebral blood volume (CBV) was varied by phantom slice, mean transit time (MTT) by column, delay time (T0) by row, and cerebral blood flow (CBF) by column and slice (dependent on CBV and MTT), and Tmax by row and column (dependent on T0 and MTT).

### Derivation of the Calibration Relationship

Let Equation (9) be the linear regression of the model-independent (MI) estimate of blood flow, *F*_*MI*_ against true blood flow, *F*_*GT*_, in the digital perfusion phantom:

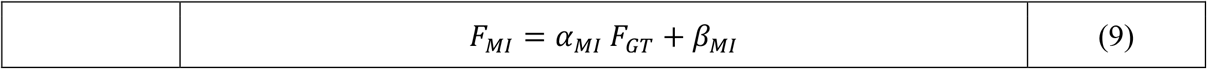

where *α* and *β* are the slope and intercept of the linear regression, respectively. Similarly, let Equation (10) be the linear regression of the model-based (MB) estimate of blood flow, *F*_*MB*_ against *F*_*GT*_ in the digital perfusion phantom:

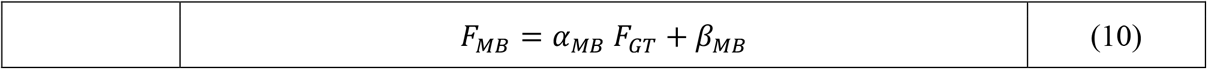

The calibration relationship for predicting the *F*_*MB*_ that is equivalent to a *F*_*MI*_ based on linear regression against *F*_*GT*_ can be derived by expressing *F*_*MB*_ in terms of *F*_*MI*_ using Equations (9) and (10):

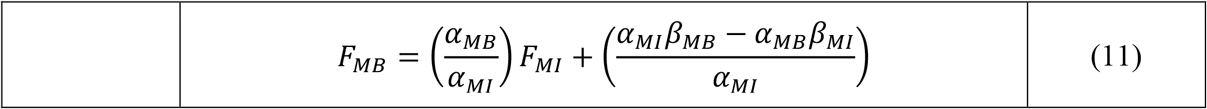

Thresholds of relative blood flow, defined as blood flow normalized by that in the normal brain hemisphere, were used identify ischemic core in this study. Let *N*_*GT*_, *N*_*MI*_, and *N*_*MB*_ be ground truth, model-independent, and model-based normal blood flow with which blood flow is normalized. *N*_*MI*_ and *N*_*MB*_ can be computed with *N*_*GT*_ and Equations (9) and (10):

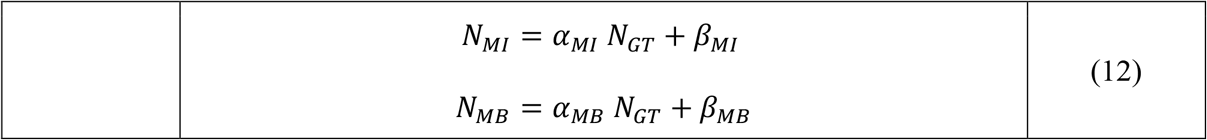

*N*_*GT*_ *=* 50 ml/min/100 g was used in this study. The model-independent and model-based relative blood flow thresholds, *R*_*MI*_ and *R*_*MB*_, respectively, were then:

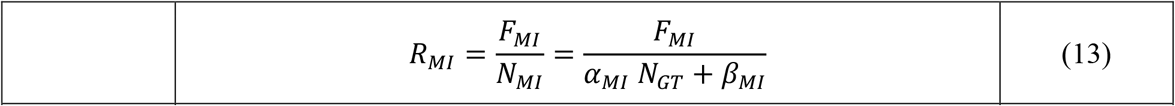

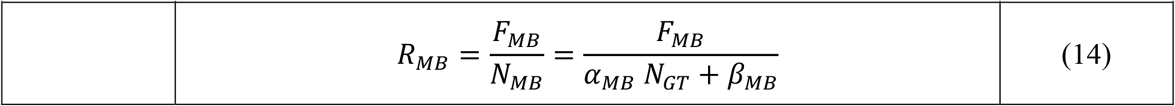

Substituting (11) into (14) and using (13) to express the result in terms of *R*_*MI*_ as shown in Equation (7) of the main text:

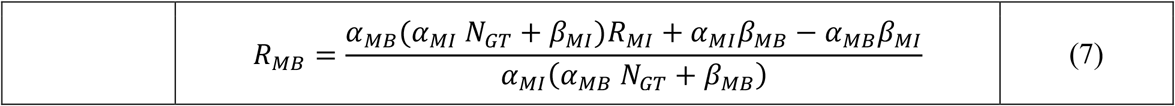

Equation (7) is the calibration relationship by which the equivalent model-based relative blood flow threshold to a reference model-independent relative blood flow threshold can be determined.

Using a similar methodology for *T*_*max*_ would yield the following calibration equation between model-independent Tmax and model-based Tmax like that of Equation (11) and as shown in Equation (8) in the main text:

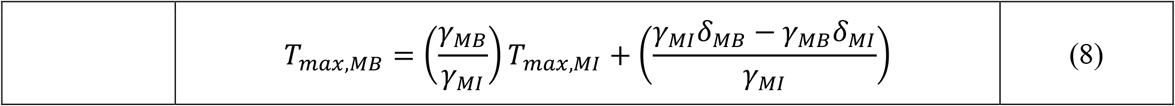

where *γ* and *δ* are the linear regression slope and intercept of estimated Tmax against ground truth Tmax in the digital perfusion phantom.

